# Mass azithromycin distribution and nutritional status among children aged 1-59 months: a secondary analysis of a cluster randomized trial

**DOI:** 10.1101/2025.08.17.25333846

**Authors:** Mamadou Ouattara, Huiyu Hu, Mamadou Bountogo, Boubacar Coulibaly, Valentin Boudo, Guillaume Compaoré, Clarisse Dah, Elodie Lebas, Kieran S. O’Brien, Benjamin F. Arnold, Thomas M. Lietman, Ali Sié, Catherine E. Oldenburg

## Abstract

**Background:** Twice yearly distribution of azithromycin has been shown to reduce all-cause child mortality in several contexts. Azithromycin likely reduces mortality via reduction in clinical or subclinical infections. Treating infections may improve child nutritional status to further reduce mortality risk. Here, we used data from a placebo-controlled cluster randomized trial of azithromycin for prevention of child mortality in Burkina Faso to evaluate the effects of twice-yearly azithromycin on mid-upper arm circumference (MUAC) in children aged 1-59 months and weight-for-age Z-score (WAZ) in infants aged 1-11 months.

**Methods:** The Child Health with Azithromycin Trial (CHAT) was a 1:1 cluster randomized trial of twice-yearly azithromycin compared to matching placebo for prevention of all-cause mortality in children aged 1-59 months. MUAC measurements were collected in all children at the time of treatment. Weight measurements were collected in infants aged 1-11 months to calculate weight-based dosing. We evaluated MUAC and WAZ at the community level and longitudinally at the individual child level, accounting for the cluster randomized design of the trial.

**Results:** 80,282 children aged 1-59 months and 26,983 infants aged 1-11 months in 284 communities were included in analyses. At the community level, we found no evidence of a difference in mean MUAC or WAZ after 36 months or over time. There was no evidence of a difference in change over time in MUAC (*P*-value for interaction = 0.94) or WAZ (*P*=0.18) at the individual level.

**Conclusions:** We found no evidence to support an effect of twice-yearly mass azithromycin distribution on measures of undernutrition in children in this setting in Burkina Faso.

**Trial Registration:** ClinicalTrials.gov NCT03676764

## INTRODUCTION

Twice yearly mass distribution of azithromycin has been shown to reduce all-cause child mortality in children aged 1-59 months in high mortality settings.^1–3^ Azithromycin is a broad-spectrum antibiotic, and this effect is likely largely due to reductions in transmission of pathogens. Previous work has suggested that azithromycin reduces mortality due to a number of infectious causes.^4^ Some studies have shown that antibiotic use increases child growth in children with pre-existing morbidity or infection.^5,6^ Individually randomized trials of azithromycin in infants without pre-existing morbidity have not shown evidence of an effect of azithromycin compared to placebo on growth endpoints.^7,8^ However, individual (rather than community-wide) distribution of azithromycin to infants has not been shown to reduce mortality, and thus there may be effects at the community level that are not observed with individual dosing.^9,10^

Studies of mass distribution of azithromycin for child survival and trachoma have not found evidence of an effect of azithromycin for child nutritional status.^11–14^ These studies have generally evaluated nutritional status at a single time point or longitudinally several years apart. Here, we used data from a cluster randomized trial in Burkina Faso that collected mid-upper arm circumference (MUAC) measurements on all children and weight measurements on all infants <12 months of age to evaluate the impact of twice-yearly distribution of azithromycin to children aged 1-59 months on both point prevalence of undernutrition and longitudinal growth in children with repeated anthropometric measurements compared to placebo. We hypothesized that anthropometric indicators of nutritional status would be higher in communities receiving twice-yearly azithromycin compared to placebo.

## METHODS

### Trial methods

The Child Health with Azithromycin Treatment (CHAT) trial was a cluster randomized placebo-controlled trial evaluating the effect of twice-yearly mass azithromycin distribution compared to placebo for prevention of child mortality.^1,15^ The study was conducted from August 2019 through February 2023. The primary outcome, all-cause mortality among children aged 1-59 months, has been previously reported and was consistent with an 18% reduction in mortality in communities receiving twice yearly azithromycin for 36 months compared to placebo.^1^ Here, we report non-prespecified anthropometric outcomes. The trial was reviewed and approved by the Institutional Review Board at the University of California, San Francisco and the Comité National d’Ethique pour la Recherche in Ouagadougou, Burkina Faso. Written informed consent was obtained from the caregiver of each participant.

### Study setting

The CHAT trial was conducted in Nouna District in northwestern Burkina Faso. The district is rural and largely agrarian, with seasonal rainfall from June through September and an annual harvest in November.^16^ Undernutrition is common in Nouna, with a wasting prevalence (weight-for-height Z-score, WHZ, < −2) reported at 6% and stunting prevalence (height-for-age Z-score, HAZ) of 24% in 2017-2019.^17^ All communities in the district were eligible to participate, however an escalating insecurity situation over the course of the trial meant that some communities were no longer safe to access by the study team. Communities that were inaccessible were eligible to re-enter the study if it became safe for them to do so.

### Communities and Participants

Prior to the first study census, a census of all communities and structures in the district was undertaken. All communities that agreed to participate in the study were eligible for participation and were randomized, with the exception of the town of Nouna, which was excluded because it is more urban and has lower mortality rates.

Communities with a census population of 2000 or more were split into multiple randomization units (“clusters”), so each cluster had a population of <2000 individuals. During the study, a census was conducted twice yearly to enumerate all children in the study area. Study treatment was provided during the census. Children were eligible for treatment if they were between 1 and 59 months of age, had no documented allergies to macrolides, and the caregiver consented to their participation in the study.

### Randomization, interventions, and masking

Communities were randomized in a 1:1 fashion to twice-yearly azithromycin distribution or placebo to children aged 1-59 months. Communities in the azithromycin group received a single, oral 20 mg/kg dose of azithromycin delivered to all children aged 1-59 months, with the dosing based on weight for children < 12 months of age or based on height stick approximation for children ≥ 12 months.^18,19^ Communities in the placebo arm received an equivalent volume of matching placebo. The placebo was identical to the azithromycin with the active ingredient to facilitate masking. Families, study staff, and investigators were masked to treatment allocation.

### Anthropometric measurements

Weight measurements were taken among infants < 12 months of age to facilitate dosing at each biannual census prior to treatment administration. MUAC measurements were collected on all children regardless of age. Children with MUAC measurements < 11.5 cm were referred to care for severe acute malnutrition (SAM) care. Children were linked with a unique identification number across multiple time points as part of the census. We used longitudinal measurements to assess changes in MUAC over time at the individual level in children and 6-month changes in WAZ in the subset of infants with two measurements. We also evaluated repeat cross sectional anthropometric measures at the community level for both MUAC and WAZ.

### Statistical analysis

This was a non-prespecified secondary analysis of the CHAT trial.^1^ The sample size for CHAT was based on the primary outcome, all-cause child mortality, with a fixed sample size of 341 clusters based on the number of clusters identified during study planning.

### Community level analysis

We summarized mean MUAC and WAZ at each census round by cluster, then aggregated these to estimate arm-specific means and 95% confidence intervals over time. We also plotted the mean MUAC and WAZ trajectories across study phases by treatment arm to visualize temporal trends. In addition, we fitted linear mixed-effects models on mean cluster-level MUAC and WAZ, including treatment arm, study phase, and their interaction, with random intercepts for clusters to account for repeated measures.

### Individual-level analysis

We fitted linear mixed-effects models to account for repeated measurements within children and clustering by community. For MUAC, the model included treatment arm, census round, child age at visit, and baseline MUAC, with random intercepts for children nested within clusters to account for the longitudinal structure of the data. For WAZ, which is inherently age-standardized, we fitted models with treatment arm and baseline WAZ as covariates. Because weight was only measured for children aged 1–11 months, resulting in two timepoints, we included random intercepts only at the cluster level. Baseline age was restricted to children younger than 6 months. As a sensitivity analysis, we further explored potential effect modification by age on both MUAC and WAZ outcomes by including an age-by-treatment interaction term. All analyses were conducted in R (version 4.4).

## RESULTS

Of 341 communities randomized to twice yearly azithromycin or placebo, 284 entered into the study during the first census (N=144 azithromycin, N=140 placebo, **Figure 1**). A total of 80,282 children and 26,983 infants were included in community-level analyses for MUAC and WAZ outcomes, respectively. A total of 64,762 children had longitudinal MUAC measurements and 10,271 infants had longitudinal WAZ measurements and contributed to individual-level analyses. Baseline characteristics were similar in azithromycin and placebo-treated children (**Table 1**).

**Table 1.** Baseline demographic characteristics.

|  | **Azithromycin** | **Placebo** |
| --- | --- | --- |
| Number of communities | 144 | 140 |
| Number of children (1-11 months) with WAZ | 3,206 | 3,204 |
| Number of children (1-59 months) with MUAC | 19,986 | 20,261 |
| Child's sex among children aged 1-11 months, N (%) |  |  |
| Female | 1,516 (47.3%) | 1,598 (49.9%) |
| Male | 1,690 (52.7%) | 1,606 (50.1%) |
| Child's sex among children aged 1-59 months, N (%) |  |  |
| Female | 9,837 (49.2%) | 10,059 (49.6%) |
| Male | 10,149 (50.8%) | 10,202 (50.4%) |
| Age among children aged 1-11 months, month, median (IQR) | 6 (3 to 8) | 6 (3 to 8) |
| Age among children aged 1-59 months, month, median (IQR) | 34 (20 to 47) | 34 (20 to 47) |
| WAZ, median (IQR) | -0.92 (-1.86 to -0.01) | -0.9 (-1.9 to 0.03) |
| MUAC, cm, median (IQR) | 14.6 (14.0 to 15.5) | 14.5 (13.8 to 15.3) |

Abbreviations: WAZ, Weight-for-Age z-score; MUAC, Mid-Upper Arm Circumference; IQR, interquartile range

**Figure 1.**
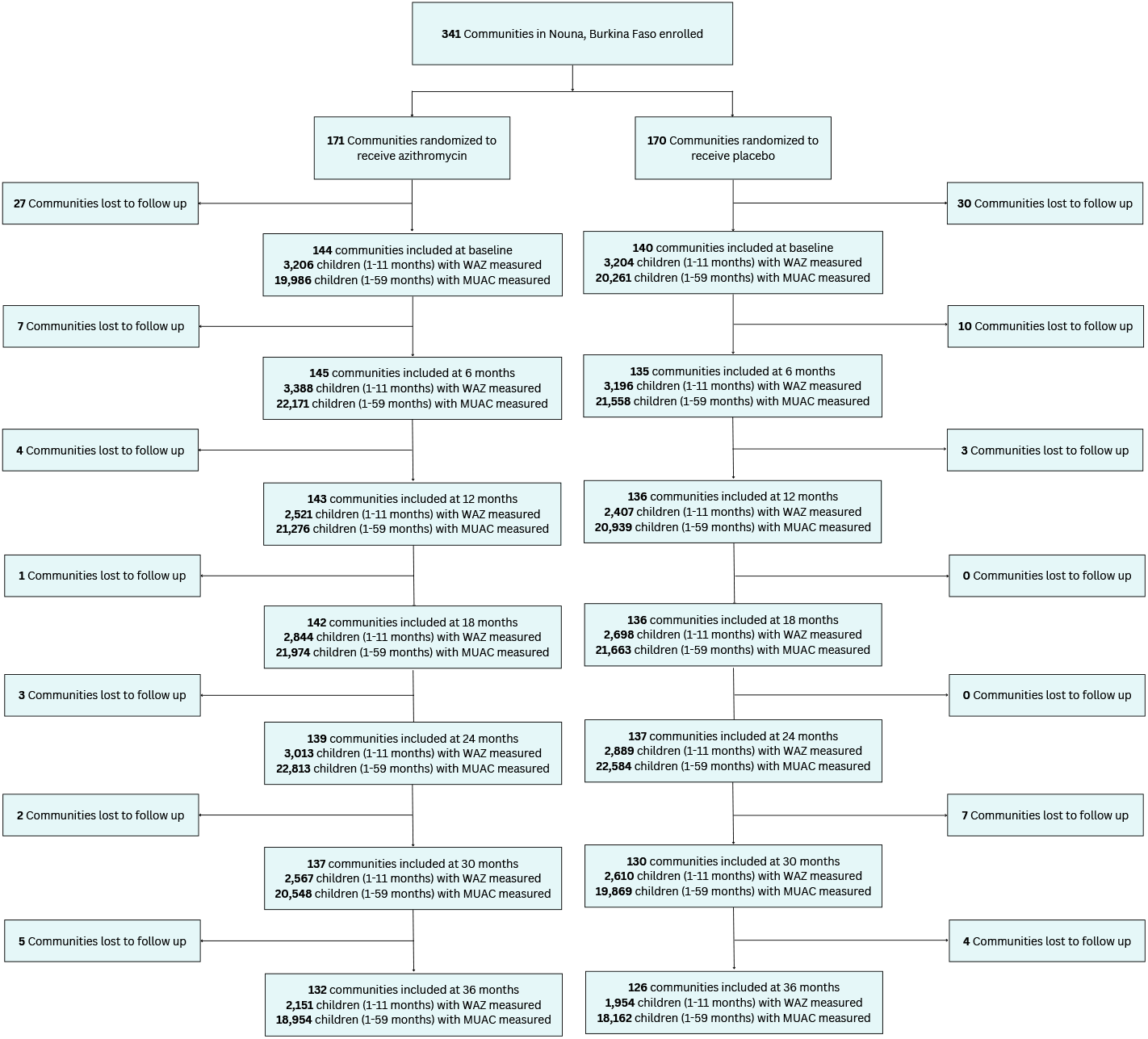
CONSORT flow diagram of included communities and participants.

At the community level, we found no evidence of a difference in mean MUAC or WAZ 36 months or at any interim time point (**Figure 2**). At 36 months, mean MUAC was 14.7 cm in each, with a difference of −0.01 cm (95% CI: −0.13 to 0.11; p=0.90). The mean WAZ at 36 months was −0.81 in the azithromycin group and −0.95 in the placebo group, yielding a difference of −0.14 (95% CI: −0.41 to 0.13; p=0.30). There was no evidence of a difference in MUAC or WAZ over time (overall treatment arm by time P-value = 0.18 for WAZ and 0.19 for WAZ).

**Figure 2.**
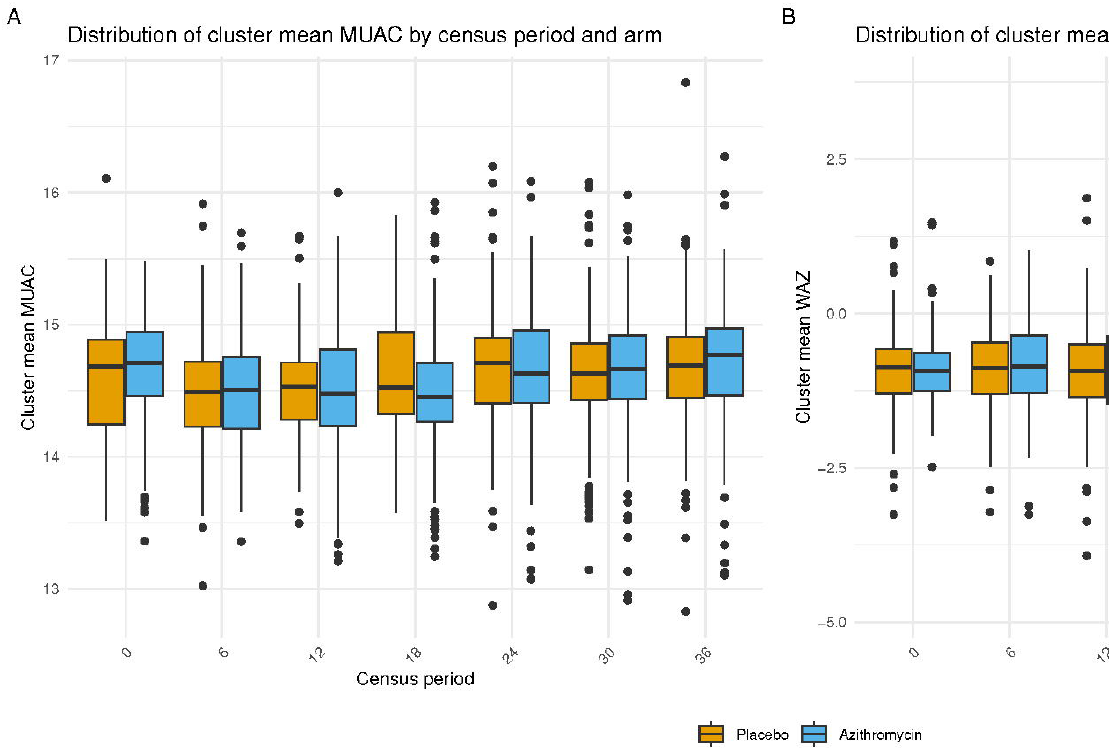
Anthropometric measurements by treatment group over time at the cluster level. **A.** Mean mid-upper arm circumference (MUAC) in children aged 1-59 months. **B**. Weight-for-age Z-score (WAZ) in children aged 1-11 months.

At the individual level, MUAC increased over time in both groups but there was no evidence of a difference in MUAC by group over time (P-value for interaction = 0.94; **Figure 3**). Overall, younger children were more likely to have increased MUAC if they were in the azithromycin arm compared to placebo compared to older children (*P*-value for interaction < 0.001), but the 95% confidence interval included 0 (no effect) for the difference in MUAC in azithromycin compared to placebo communities across all age groups (**Supplemental Figure 1**). In analyses of WAZ in infants with two WAZ measurements (i.e., infants < 6 months at the first measurement), WAZ was −1.1 (95% CI −1.2 to −1) in infants in azithromycin communities compared to −1.2 in placebo communities (95% CI −1.3 to −1.1). This corresponded to a mean difference of 0.08 (95% CI −0.04 to 0.21, *P*=0.18) in azithromycin compared to placebo communities (**Figure 3**). The effect of azithromycin compared to placebo increased with age among infants aged 1 to 5 months at their first measurement, but the P-value for interaction was not statistically significant (*P*=0.12; **Supplemental Figure 1**).

**Figure 3.**
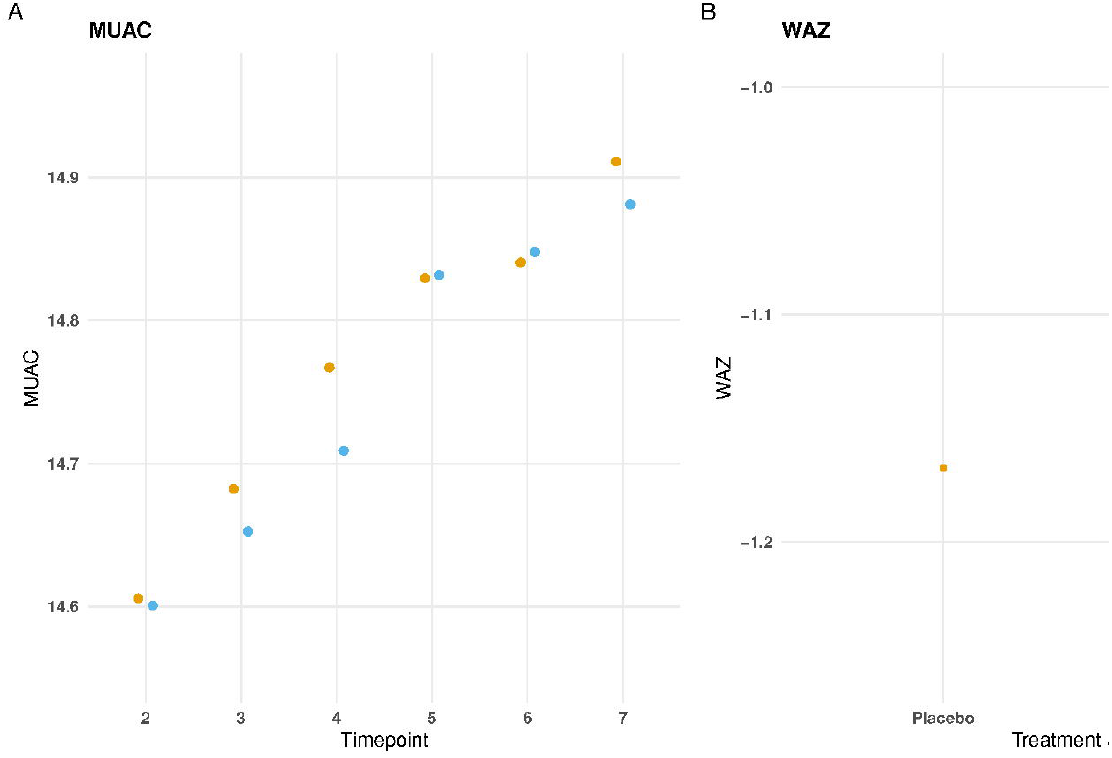
Individual-level analyses of azithromycin compared to placebo. **A.** Mid-upper arm circumference (MUAC) over time in children aged 1-59 months with >1 MUAC measurement; **B**. Weight-for-age Z-score (WAZ) in infants age 1-5 months at the first measurement with one additional WAZ measurement. Panel B depicts mean WAZ with 95% confidence intervals at the second measurement (6 months later) in the azithromycin compared to placebo group. For both panels, yellow indicates the placebo and blue indicates azithromycin group.

## DISCUSSION

In this non-prespecified secondary analysis, we found no evidence to support an effect of twice-yearly azithromycin distribution on anthropometric measurements at the community or individual level after 36 months of distributions. Although we hypothesized that mass azithromycin distributions may improve nutritional status by reducing clinical or subclinical infections (e.g., due to enteric infections)^4,20^, there was no evidence of an effect on MUAC in children under age 5 or WAZ in infants under 12 months of age. These results are consistent with previous studies which have failed to show an effect of individual or mass distribution of azithromycin on child growth.^5,7,8,12–14,21^ In contrast, a meta-analysis of trials of antibiotic use in children with morbidity (e.g., HIV or malnutrition) suggested that antibiotics may be growth promoting in these children.^5^ The results of this meta-analysis were largely driven by a trial of antibiotics for uncomplicated severe acute malnutrition.^22^ Antibiotics are included as part of routine treatment for uncomplicated severe acute malnutrition for children without clinical signs of infection due to the potential for asymptomatic infection in these children. In general, antibiotic trials in this population have shown short-term (up to 8 weeks) but not longer-term effects on weight gain.^23,24^ It is possible that azithromycin has a similar short-term effect in well-nourished children that was not captured in the present study due to the timing of measurements. However, these results suggest that growth promotion is unlikely to be a major contributing mechanism for the child survival benefits of mass azithromycin distribution.

Because these data were collected as part of a large, simple randomized controlled trial, data collection was limited to MUAC in all children and weight in infants due to logistics and feasibility in transporting anthropometric equipment to the field. As a result, height was not collected and thus we were unable to evaluate the impact of azithromycin in linear growth. Stunting, or a low height-for-age Z-score (HAZ), reflects chronic undernutrition and is an important indicator for monitoring malnutrition. Antibiotic trials have generally shown increased impact of antibiotics on weight gain rather than linear growth^5^, perhaps due to the antibiotic’s shorter-term but not longer-term effects. Whether repeated dosing at regular intervals with an antibiotic, such as in mass drug administration programs that may occur for several years of a child’s life, affects linear growth differently than individual shorter duration dosing is unknown.

The results of this study must be interpreted in the context of several limitations. As previously mentioned, measurements were collected at 6-month intervals that would not capture any effect of antibiotics on shorter-term growth that did not persist to 6 months. Weight measurements were only collected in children < 12 months of age, as they were collected as part of weight-based dosing for infants. These measurements were collected for the purposes of calculating the dose of azithromycin or placebo using hanging infant scales, which may have introduced measurement error. We anticipate that any measurement error in weight measurements for infants would be non-differential with respect to azithromycin or placebo, due to the masked nature of the trial, and thus would attenuate effects towards the null. This study evaluated twice-yearly distribution of single-dose azithromycin in the general population of children aged 1-59 months. While there is some evidence that the effects of azithromycin for mortality are stronger in children with undernutrition^25–27^, current evidence does not support targeted azithromycin distribution for mortality. Whether different dosing strategies, timing, or duration would yield different effects is not known. Finally, the results of this study can likely only be generalized to settings with a similar distribution of malnutrition, food insecurity, dietary practices, and background antibiotic use.

Overall, these results do not support an effect of twice-yearly azithromycin distribution on MUAC in children aged 1-59 months or WAZ in infants aged 1-11 months 6 months after distribution.

Changes in ponderal growth are unlikely to be a major contributor to the protective effect of azithromycin for child mortality in Burkina Faso.

## Data Availability

All data produced in the present study are available upon reasonable request to the authors

